# Charting the Circulating Proteome in ME/CFS: Cross System Profiling and Mechanistic insights

**DOI:** 10.1101/2025.05.28.25328245

**Authors:** August Hoel, Fredrik Hoel, Sissel Elisabeth Furesund Dyrstad, Henrique Chapola, Ingrid Gurvin Rekeland, Kristin Risa, Kine Alme, Kari Sørland, Karl Albert Brokstad, Hans-Peter Marti, Olav Mella, Øystein Fluge, Karl Johan Tronstad

**Author notes:** These authors contributed equally (August Hoel, Fredrik Hoel).

## Abstract

Myalgic Encephalomyelitis/Chronic Fatigue Syndrome (ME/CFS) is a debilitating condition often triggered by infections. The underlying mechanism remains poorly understood, and diagnostic markers and effective treatments are presently lacking. We performed aptamer-based serum proteomics in 54 ME/CFS patients and 27 healthy controls and identified 1823 of 7326 aptamers reporting differences between the groups (845 after false discovery rate (FDR) correction). Distinct patterns of tissue- and process-specific changes were seen. There was a broad increase in secreted proteins, while intracellular proteins, e.g. from skeletal muscle, particularly showed reduction. Immune cell-specific signatures indicated immune reprogramming, including a distinct reduction in neutrophil-associated proteins. Focused secretome analysis supported intensified regulatory interactions related to immune activity, inflammation, vasculature, and metabolism. Validation of measurements using antibody-based methods confirmed findings for a selection of proteins. The uncovered serum proteome patterns in ME/CFS patients help clarify a multifaceted pathophysiology and offer a foundation for future therapy and biomarker discovery efforts.

## Introduction

Myalgic Encephalomyelitis/Chronic Fatigue Syndrome (ME/CFS) is a debilitating disease that often is linked to infection. Common triggers include Epstein–Barr virus (EBV, infectious mononucleosis), human herpes virus, and more recently, SARS-CoV-2 (COVID-19) ^1–3^. Using the Canadian consensus criteria ^4^, the pre-COVID-19 pandemic prevalence of ME/CFS was estimated to be between 0.2% and 0.8% ^5–7^. There has been an increase in cases following SARS-CoV-2 ^8^. Key symptoms include profound fatigue, post-exertional malaise (PEM), sensory hypersensitivity, pain, unrefreshing sleep, and cognitive dysfunction ^9,10^. The consequences of ME/CFS are severe for both patients and their families, with high socio-economic costs ^7,11–13^. There is an urgent need for more knowledge about the biological mechanisms of ME/CFS to develop effective diagnostic markers and treatments.

Although the etiology of ME/CFS remains unclear, several biological manifestations, such as immune system dysregulation, viral persistence, chronic inflammation, and impaired metabolism, have been reported ^14,15^. We are currently investigating the hypothesis that ME/CFS may involve an autoimmune mechanism leading to vascular dysregulation, causing tissue hypoperfusion and hypoxia, especially upon exertion, and resulting in both short- and long-term effects on energy metabolism ^16,17^. This mechanism could explain key aspects of symptom generation and fatiguability in ME/CFS, potentially involving exertion-triggered muscle abnormalities, as recently reported in association with PEM in long COVID patients ^18^.

Human cells express thousands of proteins in a tissue-specific manner ^19,20^, many of which are released into the blood either by function, as messengers, or due to tissue leak ^21^. Pathology in specific parts of the body influences both the patterns of protein expression and the release of proteins into the bloodstream through controlled and uncontrolled mechanisms. Knowledge about circulating protein levels can provide insights into disease mechanisms, tissue regulation, homeostasis, secretory processes, and metabolism ^20^. Studies of circulating proteins in the blood of ME/CFS patients have reported abnormal, yet variable, findings related to cytokines and immunological factors ^22,23^, vascular regulators ^24^, and metabolic and muscle-derived messengers ^17,25^. Further in-depth studies are needed to better understand the underlying pathology.

Aptamer microarray proteomics is a discovery platform capable of simultaneously measuring a high number of proteins from small sample volumes, which utilize the protein-specific binding affinity of small single-stranded DNA or RNA molecules (aptamers) ^26^. This technology has been used successfully to study circulatory factors involved in aging ^27^, kidney disease ^26^, diabetes ^28^, and other diseases. To our knowledge, it has been employed in only two previous ME/CFS studies, both using relatively small study cohorts ^29,30^.

The overall aim of this study was to identify circulating protein patterns that reveal new insights into processes taking place at cellular, tissue and systemic levels in ME/CFS patients, and investigate the hypothesis of an immune system-related root-mechanism causing impaired microcirculation and energy metabolism.

## Results

### Characterization of the subjects

We analyzed blood serum from 54 ME/CFS patient biobank samples collected at baseline in two previous clinical intervention trials: RituxME (ClinicalTrials.gov NCT02229942, 2014–2017) ^31^ and CycloME (ClinicalTrials.gov NCT02444091, 2015–2020) ^32^ (Table 1, Figure 1A). The healthy control (HC) group consisted of 27 sex- and age-matched individuals. The blood glucose concentration was not statistically different between the ME/CFS group and the HC group (Table 1). However, the blood concentrations of triacylglycerols (TAGs) and non-esterified fatty acids (NEFAs) were higher in the ME/CFS group. This finding is consistent with our previous comprehensive metabolomics study using samples from the same biobank, which identified patterns of energy strain and context-dependent metabolic phenotypes (metabotypes) in patient subsets ^17^.

**Table 1.** Group characteristics.

| Study groups | HC | ME | P-value |
| --- | --- | --- | --- |
| Subjects, N (%) | 29 (100) | 50 (100) |  |
| Women, N (%) | 21 (72) | 40 (80) |  |
| Men, N (%) | 8 (28) | 10 (20) |  |
| RituxME | NA | 15 |  |
| CycloME – Response | NA | 21 |  |
| CycloME – No response | NA | 14 |  |
| Fasting, N (%) | 0 (0) | 11 (22) |  |
| BMI (mean ± SD) | 24.0±2.6<sup>a</sup> | 24.4±4.2 | 0.6495 |
| Age | 38.4±9.3 | 40.4±10.1 | 0.3615 |
| Infection, N (%) | NA | 38 (76) |  |
| Mean steps 24h (mean ± SD) | NA | 3021±2001 | |
| SF36-PF (mean ± SD) | NA | 31.2±19.3 | |
| Self rep. Funct (% , mean ± SD) | NA | 16.6±7.7 | |
| Glucose (mM, mean ± SD) | 5.04±0.40 | 5.24±0.82 | 0.3828 |
| TAG (mM, mean ± SD) | 0.87±0.31 | 1.35±0.89 | 0.0254* |
| NEFA (mM, mean ± SD) | 0.21±0.13 | 0.40±0.28 | 0.0262* |

Overview of the two groups of healthy control subjects (HC) and ME/CFS patients. The ME/CFS samples were obtained from participants in the RituxME and CycloME trials. Measurements of glucose, triglycerides (TAG) and non-esterified fatty acids (NEFA) were performed in the clinic.
\*P < 0.05, welch's test; <sup>a</sup> HC BMI N=24

**Figure 1.**
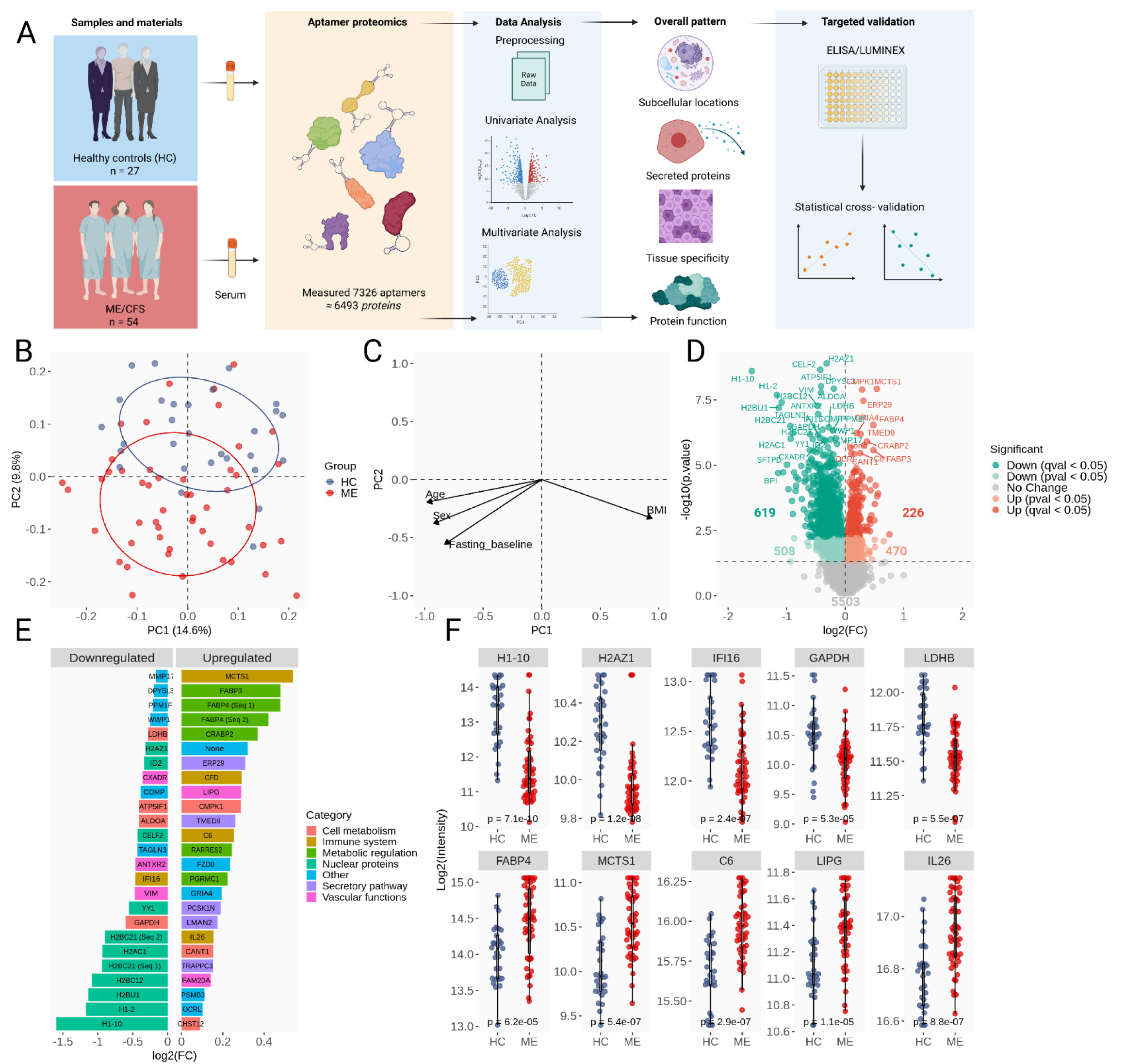
Serum proteomics comparing ME/CFS (ME) patients with healthy controls (HC) A) Study Approach: The serum protein concentrations were measured using aptamer-based technology (SomaScan). Exploratory data analysis (EDA) was conducted to identify cellular and tissue-specific impacts, effects on the secretome, and the associated molecular and biological functions involved. Antibody-based measurements were used to validate and expand on findings, with a focus on the immune system and energy metabolism. B) Principal Component Analysis (PCA) with overlay of the ME/CFS and HC groups, and C) projections of associated eigenvalues reflecting the influence of age, sex, BMI, and fasting on the PC1 and PC2 dimensions. D) Volcano plot displaying all the p- and q-significant features. E) The 25 most up- or downregulated protein targets (ranked by fold change) in the ME/CFS patients are categorized according to their functional roles (colors), based on current literature. F) Group comparison of selected significantly affected serum proteins. All statistical outcomes include both uncorrected (p-value) and multiple comparison corrected (FDR, q-value) univariate analyses, adjusted for sex, age, BMI, and overnight fasting.

### Comprehensive differences between the ME/CFS group and the HC group

The serum samples were analyzed using the SomaScan aptamer-based detection platform, yielding a dataset for 7326 aptamers (6494 protein targets, 6409 identified proteins) (SupplData1). In total, as many as 1823 of the aptamers showed statistically significant differences (p < 0.05) between the ME/CFS group and the HC group, with 845 showing q-significant changes (SupplData2). This corresponds to 1610 different proteins (751 q-significant) demonstrating altered serum concentrations.

The PCA (Principal Component Analysis) plot with group overlay showed partial separation between the ME/CFS and HC subjects (Figure 1B-C; SupplData3), primarily along the PC2 dimension (y-axis, 9.6% explained variance). The groups largely overlapped across the PC1 dimension (x-axis, 14.5% explained variance), indicating that certain elements of intragroup variation are shared between the groups. This makes sense, as PC1 was more influenced by the covariate factors, age, sex, BMI, and fasting state, as indicated by the respective loadings, compared to PC2 (Figure 1C; variable, PC1, PC2, p; BMI, 0.943 -0.333, 0.106; age, -0.981, -0.195, 0.381; sex, -0.925, -0.380, 0.091; fasting, -0.830, -0.557, 0.046). Since group separation along PC2 did not seem to be driven by these covariate factors, this may point to more disease-specific changes. Accordingly, the major PC2-driving proteins (i.e. with high loading) expressed more statistically significant differences between the two groups, compared to major PC1-driving proteins. Additional investigations aimed at separating disease-specific effects from covariate influences are included in the forthcoming analyses.

More than 60% of the affected aptamers showed lower protein abundance in the ME/CFS patients (62% using p < 0.05, 1127/1823; 73% using q < 0.05, 619/845), while the remainder showed higher abundance (38% using p < 0.05, 696/1823; 27% using q < 0.05, 226/845) (Figure 1D).

### Initial key observations related to the hypothesized pathomechanism

The list of the 25 protein targets showing the sharpest increase or decrease in ME/CFS patients indicated specific changes of key interest (Figure 1E). The proteins that showed the most decline were predominantly intracellular proteins, such as histones from the nucleus and enzymes involved in metabolic pathways. In contrast, many proteins involved in secretory and systemic regulation of the immune system and metabolism showed significant increase. Additionally, several factors related to vascular function were among the most affected proteins, either increased or decreased. Individual data are shown for a selection of the most significantly affected proteins (Figure 1F).

Among the proteins that showed the most comprehensive reduction were a group of histones (H1x, H2.1, and several H2B and H2A types) (Figure 1E-F). These are normally nuclear proteins but can be released by activated neutrophils as part of neutrophil extracellular traps (NETs)^33^. Other proteins showing prominent decreases included several enzymes related to cellular energy metabolism, such as glyceraldehyde-3-phosphate dehydrogenase (GAPDH), fructose-bisphosphate aldolase A (ALDOA), and L-lactate dehydrogenase B chain (LDHB). We also observed significant reductions in several proteins associated with vascular function and responses to cellular oxygen deprivation, including vimentin (VIM) ^34^, anthrax toxin receptor 2 (ANTXR2, also known as capillary morphogenesis gene 2 (CMG2)) ^35^, coxsackievirus and adenovirus receptor (CXADR) ^36^, and ATP synthase inhibitory factor subunit 1 (ATP5IF1) ^37^. Other reduced proteins included the neuron protein Transgelin-3 (TAGLN3), gamma-interferon-inducible protein 6 (IFI16), and cartilage oligomeric matrix protein (COMP).

The proteins showing the greatest increase in the ME/CFS patients included several factors associated with immune responses (Figure 1D-F), such as malignant T-cell-amplified sequence 1 (MCTS1) ^38,39^, complement factor D (CFD), complement component C6 (C6), and the cytokine IL26. There were also significant elevations in factors involved in metabolic regulation, including fatty acid-binding proteins 3 and 4 (FABP3, FABP4), which are considered metabolic stress factors when released into the blood ^40^. Furthermore, there were significant increases in cellular retinoic acid-binding protein 2 (CRABP2), a member of the fatty acid-binding protein family; the adipokine and immune cell chemoattractant retinoic acid responder protein 2 (RARRES2; chemerin) ^41^; and endothelial cell-derived lipase (LIPG), which is involved in lipoprotein metabolism and vascular biology ^42^ as well as immune activation^43^. Other increased proteins included endoplasmic reticulum resident protein 29 (ERP29), transmembrane emp24 domain-containing protein 9 (TMED9), pseudokinase FAM20A, which may function in hematopoiesis, and soluble calcium-activated nucleotidase 1 (CANT1).

Throughout our exploration of the data, we did not identify what we regard as statistically and mechanistically supported biomarker candidates. Yet we believe that the findings may shed light on biological mechanisms that can be targeted for such purposes.

### Influence by covariates and lack of physical activity

Covariates such as sex, age, BMI, and diet have significant impact on the composition of circulatory proteins in blood. Furthermore, in ME/CFS and other chronic diseases, the lack of physical activity (deconditioning) may cause systemic effects that influence the levels of blood protein. To evaluate the influence of sex, age, BMI, and overnight fasting on the affected proteins, we investigated the percentage of variance explained by each of these covariates compared to the ME/CFS diagnosis (Figure 2A, SupplData4). The 30 protein targets with the highest variance explained were selected for each covariate in the linear regression model. Overall, the proteins highly influenced by the ME/CFS diagnosis showed relatively small additional influence from the tested covariates, which suggest that these are disease-specific alterations. This included several of the aforementioned proteins, such as histones (H2AZ1, H1-10, and others), MCTS1, VIM, and LDH. In comparison, the covariates showed associations with separate, yet partially shared, sets of proteins. Among the proteins most affected by several covariates were metabolic hormones, such as leptin, FABP3, and FABP4. Altogether, this analysis identified protein variation caused by sex, age, BMI, and overnight fasting, and separated this from protein alterations specifically observed in ME/CFS patients.

**Figure 2.**
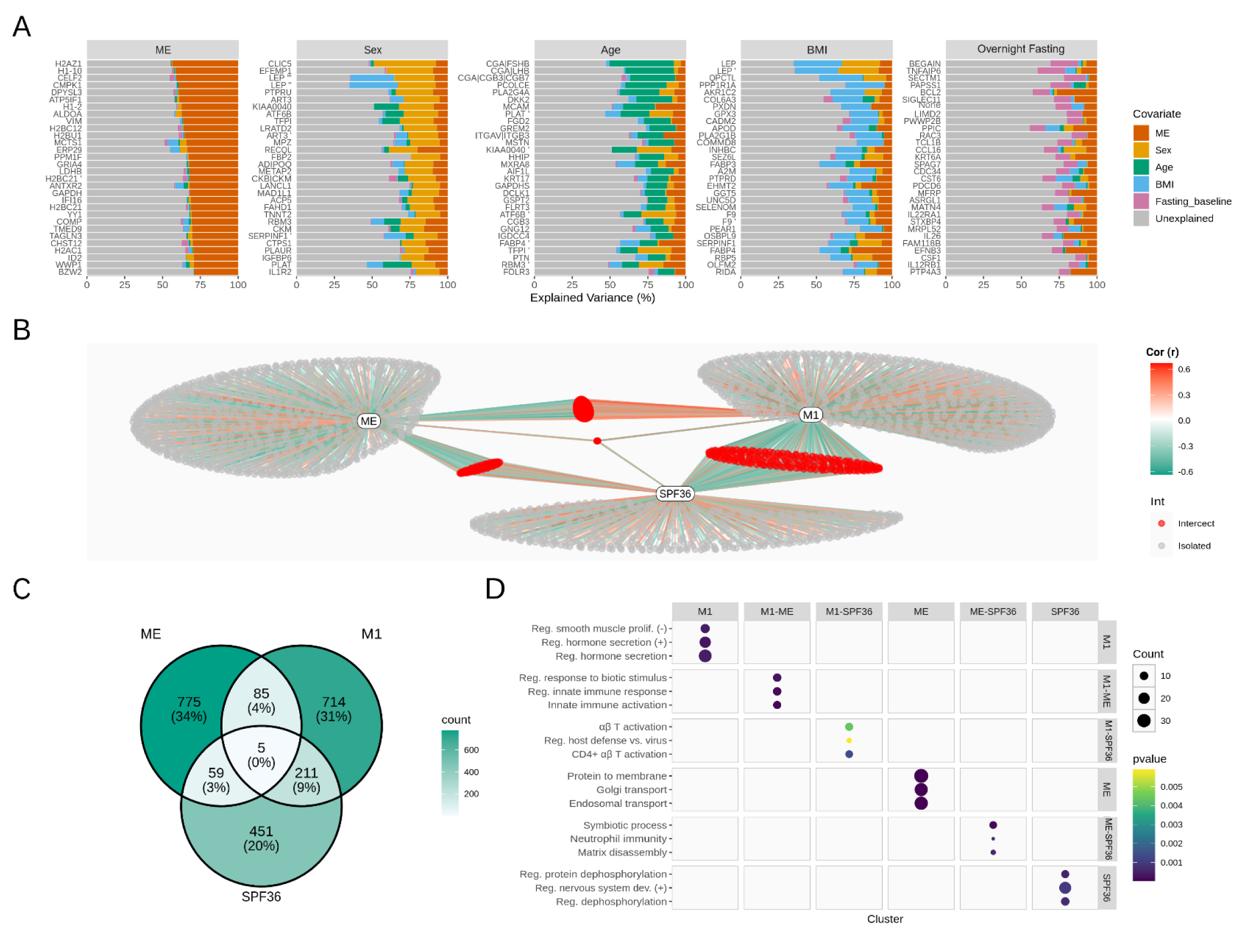
Analysis of influence by covariate factors. A) Analysis of differentially expressed features, showing the proportion of variance explained by each covariate (ME, sex, age, BMI, and fasting state). The top 30 most explained features per covariate are shown. B) Semi-partial Pearson correlation network displaying associations between molecular features and ME/CFS diagnosis (ME), metabotype (represented by metabotype M1), and SF-36 physical function (SPF36), adjusted for sex, BMI, overnight fasting, and age. Color intensity indicate correlation strength (red = positive, green = negative). C) Venn diagram displaying the overlap of changed features related to ME/CFS diagnosis, metabotype (M1) and physical function (SF-36PF score, SPF36). D) Go Ontology Enrichment analysis of biological processes within network-derived feature clusters. Dot size represents protein count, and color indicates significance (p-value).

To evaluate the extent of changes associated with physical function level and/or metabolic adaptations secondary to the ME/CFS diagnosis, we used a semi-partial Pearson correlation analysis adjusted for the covariates. This analysis identified associations between serum protein concentrations and the ME/CFS diagnosis, the metabolic phenotype (metabotype; we used metabotype M1 as reference ^17^), and the SF-36 physical function score (SF-36PF) (Figure 2B-D, SupplData5). Using the entire cohort of ME/CFS and HC individuals, a total of 924 aptamers were significantly associated with the ME/CFS diagnosis. Of these, 775 were only associated with the ME/CFS diagnosis and not influenced by the metabotype nor the SF-36PF score. Enrichment analysis indicated an impact on annotated functions such as protein localization to the plasma membrane, Golgi transport, and endosomal transport (Figure 2D). Separately, the metabotype and the SF-36PF score influenced the levels of 1015 and 726 aptamers, respectively. The 714 aptamers showing specific relationship with metabotype indicated an impact on the regulation of hormone secretion and the negative regulation of smooth muscle cell proliferation. The 451 aptamers specifically influenced by SF-36PF showed enrichment towards the regulation of protein phosphorylation and the positive regulation of nervous system development. As expected, a relatively high number of proteins (216 aptamers) were simultaneously associated with both metabotype and physical function, but only 5 of these demonstrated additional influence by the ME/CFS diagnosis. A total of 149 aptamers showed simultaneous associations between the ME/CFS diagnosis and either the metabotype category or SF-36PF, or both.

Although this analysis does not establish causality, the comprehensive changes found to be unrelated to physical activity argue against deconditioning as a driving factor for the serum proteome changes in the ME/CFS group. Instead, they indicate fundamentally distinct changes that are not explained by other known covariates, and therefore disease-specific in this context.

### Cellular proteins in circulation - release patterns

Cellular proteins enter the bloodstream through various routes, involving both passive and active release mechanisms, from the cell to the extracellular compartment. The contextual roles may include cargo transport, secretion, excretion of waste, and cell damage. We reasoned that changed patterns of proteins originating from different cellular compartments could reveal key information about the underlying mechanisms. For instance, a uniform pattern for enzymes normally found inside cells (e.g., LDH and CKM) would likely reflect events related to cell damage and tissue turnover, rather than changes in the regulation of cellular protein expression.

To investigate this notion, we used subcellular compartment annotations based on predicted protein structures available via the Human Protein Atlas (HPA). Of the overall 7326 aptamers, 53.4% (3911) were classified based on their protein targets as ‘intracellular,’ 25.9% (1900) as ‘membrane’ (including membrane-associated), and 17.6% (1292) as ‘secreted’ proteins, while 3.0% (223) remained unannotated (Figure 3A, SupplData2). Among the 1823 affected aptamers, a larger fraction (62.9%) belonged to the intracellular class, indicating a disproportionate effect on this protein class (Figure 3B). In fact, among all the aptamers that showed a decrease (1127), 74.9% (858) belonged to the intracellular protein class, whereas the membrane and secreted protein classes represented 12.8% (144) and 8.0% (90), respectively. In contrast, the protein targets that showed an increase (676) had significantly higher proportions in the membrane and secreted protein classes (30.9%, 209/676 and 26.5%, 179/676, respectively), and fewer in the intracellular class (42.6%, 288/676). Viewed differently, nearly 75% of the affected intracellular protein targets showed lower abundance in ME/CFS serum, compared to only 30-40% in the membrane and secreted protein classes, which exhibited significantly larger proportions of proteins with increased levels. In summary, there was a clear pattern of reduced release of intracellular proteins into the blood, accompanied by increased release of membrane and secreted proteins, in the ME/CFS patients.

**Figure 3.**
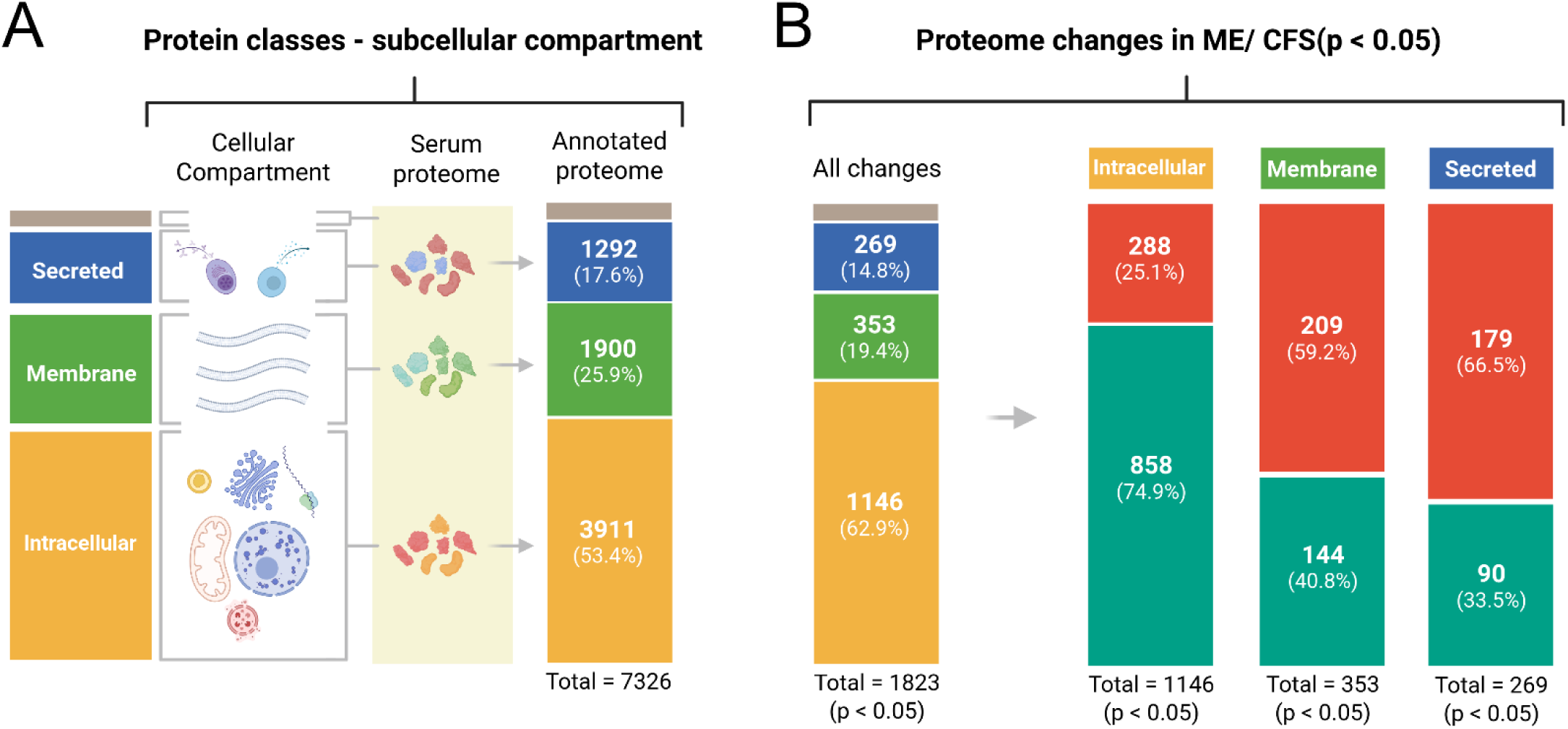
The subcellular origin of the affected proteins. The detected protein targets were classified into four categories based on their annotated subcellular localization (Human Protein Atlas): “intracellular,” “membrane” (membrane + membrane-associated), “secreted” proteins, and the remaining unannotated (“#N/A”). The accounting numbers indicate the counts and percentages of A) all the detected protein targets and B) only the affected proteins targets.

We also examined mitochondrial proteins using the MitoCarta 3.0 library ^44^. We found detectable amounts of 445 mitochondrial proteins in the serum samples (SupplData6). Approximately 20% (87) of the mitochondrial proteins showed lower blood concentrations in the ME/CFS group compared to the HC group, whereas only 7% (29) showed increase.

### Tissue and immune cell specific footprints

To evaluate the impact on major organs releasing proteins to blood, we employed tissue-specific protein panels (HPA) for the brain, liver, intestine, skeletal muscle, lymphoid tissue, and bone marrow (Figure 4A, SupplData7). Overall, a larger fraction of tissue-specific proteins from the brain, intestine, liver, and lymphoid tissue in ME/CFS patients, had increased compared rather than decreased (Figure 4B). In contrast, there was widespread reduction for proteins from skeletal muscle and bone marrow. Furthermore, when examining the subcellular origin, there were reduced levels of intracellular proteins particularly from skeletal muscle, brain, and lymphoid tissue. The pattern was different for secreted proteins, which showed primarily increased release from all the addressed tissues, except from bone marrow. The bone marrow panel showed decreased levels both for intracellular and secreted proteins in the serum from ME/CFS patients compared to the HC group. The liver and intestine panels did not indicate a clear reduction of intracellular proteins, but like most other tissues, the affected secreted proteins tended to be increased in the ME/CFS group. In summary, there was a pattern of increased secretory activity across the tissues, except for bone marrow, accompanied by reduced release of intracellular proteins from skeletal muscle, lymphoid tissue, bone marrow and brain. These coordinated tissue-level changes may support a model of an underlying root mechanism causing regulatory and adaptive systems-level responses.

**Figure 4.**
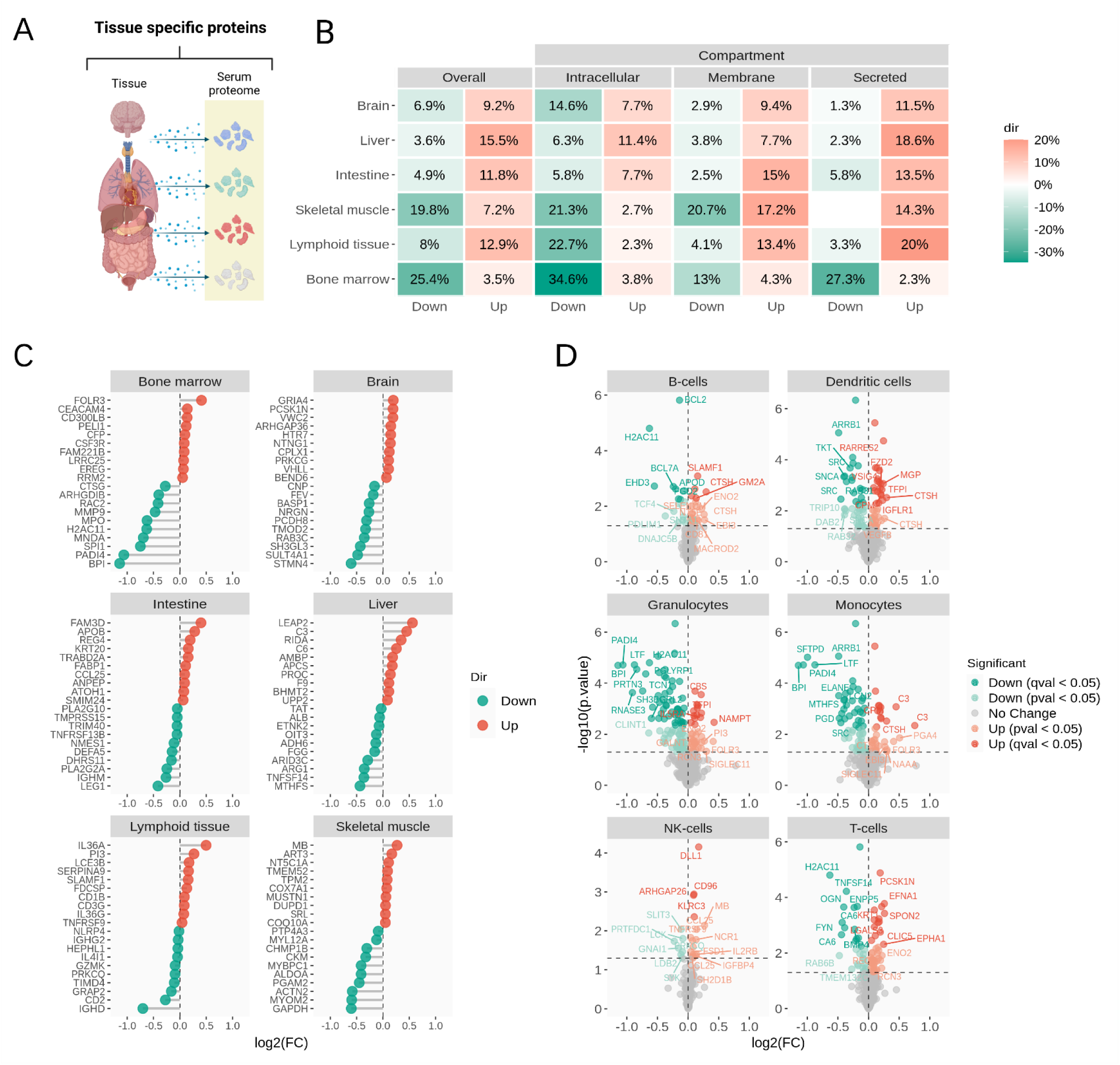
Tissue and immune cell-specific footprints. A) Tissue-related aspects were investigated using annotated protein panels specific to selected tissues and cellular compartment (HPA). B) For each protein panel, the proportion of affected proteins (up/down) was calculated relative (%) to the total number of proteins in the panel that were detected (overall), and after further division into cellular compartments. C) Plots showing the most affected proteins per tissue type (ranked by fold change). C) Volcano plots of immune cell type-specific expression panels (HPA).

More detailed investigations of the tissue-specific patterns were undertaken to obtain deeper insights into the homeostatic and functional states of affected organs and cell types. Figure 4C displays the proteins that exhibited the most significant differences between ME/CFS patients and healthy control subjects, categorized by tissue. For bone marrow proteins, proteins such as PADI4, BPI, and MPO, had a sharp reduction, which may indicate altered granulocyte/neutrophil cell function. While the brain and skeletal muscle-specific panels also showed relatively extensive reductions, the pattern was not as clear as for the bone marrow panel. Despite the lower abundance of many skeletal muscle proteins, there was a significant increase of myoglobin (MB), which may indicate a nuanced muscle pathology, rather than major muscle breakdown. For proteins from the intestine, liver, and lymphoid tissue, there was a more balanced pattern of increase and decrease. Therefore, even without detailing the specific functions of the proteins involved, ME/CFS seems to infer tissue-specific impacts on the serum proteome, potentially reflecting overall changes related to tissue homeostasis, turnover and function.

To specifically address aspects of immune dysregulation, we used annotated protein panels (HPA) specific to B-cells, T-cells, NK-cells, dendritic cells, monocytes, and granulocytes (Figure 4D). The effects associated with these cell types pointed to possible impacts on immune cell interactions, coagulation, and inflammation. As previously mentioned, multiple proteins associated with granulocytes showed significantly lower levels in the ME/CFS group compared to the HC group, such as MPO and BPI. The reduced amount of granulocyte proteins was not associated with abnormally low neutrophil counts (the most abundant type of granulocytes) or other leukocyte types in the patients (DocumentS1, Table S1). Furthermore, comparing a list of proteins associated with neutrophil granules and stimulated neutrophil protein release ^33^ ^45^, we found that about 40% or more of the proteins reported to be released by activated neutrophils showed lower serum concentrations in the ME/CFS group compared to the HC group, suggesting a suppressive effect on overall neutrophil activity.

### Secretory processes and functional changes

To further expand our investigations into the apparent stimulation of secretory processes and the impact it may infer related to ME/CFS, we utilized a revised protein-centric list of the predicted human secretome (secreted proteins) ^46^, which annotate both tissue-specific expression and functional roles of the proteins (Figure 5A, SupplData8). Across the total of 2100 aptamers targeting 1654 different secretome proteins, 454 aptamers showed significant differences between the ME/CFS group and HC group. In sharp contrast to initially classified intracellular proteins, the majority was increased (61%, 278 aptamers).

**Figure 5.**
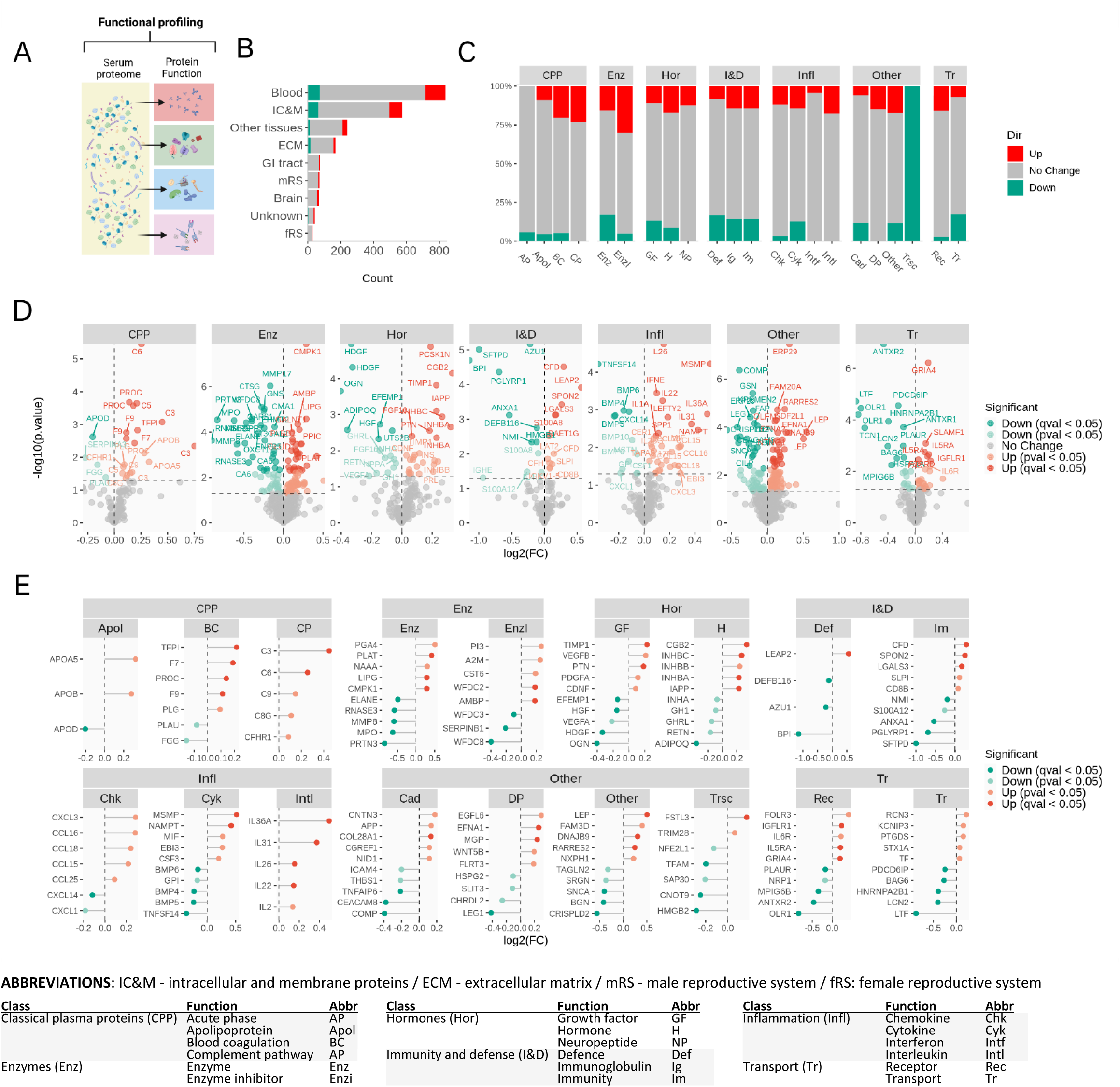
Effect on the secretome and related biological processes. A) Secreted proteins were classified according to their tissue-specificity and biological functions ^46^. B) Enrichment analysis showing the influence of ME/CFS on the nine tissue classes of secreted proteins, displaying the total and affected protein counts. C) A detailed overview of the effects on the blood protein class according to functional annotations. D) Volcano plots showing all affected secreted proteins per biological function, based on the entire secretome dataset. E) Plots showing the most affected proteins per biological function (ranked by fold change). ABBREVIATIONS: Intracellular and membrane proteins (IC&M), Extracellular matrix (ECM), Male reproductive system (mRS), Female reproductive system (fRS) - Classical plasma proteins (CPP): Acute phase (AP), Apolipoprotein (Apol), Blood coagulation (BC), Complement pathway (AP) - Enzymes (Enz): Enzyme (Enz), Inhibitor (Enzi) - Hormones (Hor): Growth factor (GF), Hormone (H), Neuropeptide (NP) - Immunity and defense (I&D): Defense (Def), Immunoglobulin (Ig), Immunity (Im) - Inflammation (Infl): Chemokine (Chk), Cytokine (Cyk), Interferon: (Intf), Interleukin (Intl) - Transport (Tr): Receptor (Rec), Transport (Tr) - Other: Cell adhesion (Cad), Transcription (Trsc), Developmental protein (DP)

The applied annotation classifies nine groups of secreted proteins according to their roles: blood proteins, proteins secreted locally in six different tissue compartments (brain, GI tract, male tissues, female tissues, unknown location, other tissues), proteins forming the extracellular matrix, and one class of intracellular and membrane-bound proteins (Figure 5B). For the extracellular matrix proteins and the intracellular and membrane-bound secretome proteins, there were relatively balanced numbers of aptamers that showed increase and decrease. In contrast, a pattern of predominant increase was seen for all the classes reflecting specific tissues (blood, brain, GI tract, male tissues, female tissues, and other tissues), indicating a systemic impact involving many organs.

The blood secretome class is crucial in the systems-level control of homeostasis, transport of nutrients, inflammatory response, defense mechanisms, hormone regulation, and many other functions. A large fraction of the secreted blood proteins is known to be released from the liver, whereas others are generated either by blood cells or more generally across all tissues. In total, 197 of 838 aptamers targeting blood secretome proteins showed significant changes, with 63% (124) showing increased concentrations and 37% (73) showing decreased concentrations. The functional annotation indicated mixed impacts related to classical plasma proteins, enzymes, hormones, immunity and defense, inflammation, and transport (Figure 5C).

To obtain a systems-level scope, functional annotation was applied to the entire secretome dataset (Figures 5D and 5E). This clearly showed a skewed pattern of increase for affected secretome proteins involved in coagulation, complement pathways, and inflammation (chemokines, interleukins). In contrast, the enzyme class showed a predominant decrease, which may relate to the overall decreased release of intracellular proteins in general (Figure 3), and especially from bone marrow (granulocytes and monocytes) (Figure 4). The other secretome classes, including hormones, immunity & defense, transport, and others, expressed mixed effects, giving a relatively balanced picture of increase and decrease, possibly reflecting different aspects of physiological regulation.

We performed a ligand-receptor interaction analysis to elucidate possible patterns of regulation that could be related to the pathomechanism of ME/CFS (DocumentS1, Figure S1). Overall, we found an overweight of cell adhesion and cytokine-cytokine receptor type ligand-receptor interactions. Interestingly, four members of the Ephrin subfamily A receptors (EPHA) and their ligands (EFNA) displayed concordance, supporting the involvement of altered EPHA-EFNA signaling in ME/CFS, as previously suggested ^29^. Furthermore, there was a positive concordance between Fibroblast growth factor receptor 3 (FGFR3) and EPHA, IL6R with the CNTF and MPZ ligands, and FLRT3 with UNC5B and UNC5D. FAP and PAM had negative concordance, along with other interactions. Based on current knowledge, it seems likely that the identified ligand-receptor interactions may play a role in ME/CFS pathology through their impact on metabolic regulation, tissue development and repair, inflammation, and angiogenesis.

### Validation and expansion of serum proteome findings using antibody-based methods

To validate and expand on findings from the aptamer-based analyses, we used antibody-based methods (ELISA and Luminex) to measure a panel of serum proteins related to immunity, inflammation, coagulation, and stressed energy metabolism. Of the 77 proteins measured on the Luminex platform (ME/CFS, n = 83; HC, n = 48), 49 proteins expressed quantifiable serum concentrations at the group level (SupplData9). There was a reasonably good correlation between the antibody-based and aptamer-based measurements, and importantly, significant effects in ME/CFS patients versus HC subjects were generally reproduced (Figures 6A-B). Thus, the selected antibody-based measurements validated key findings from the aptamer-based analyses. However, the increase in myoglobin was not reproduced by the antibody-based method. A selection of significant differences between ME/CFS and HC based on the Luminex data, are shown in the heatmap in Figure 6C, which also compare the serum protein levels according to metabotype (metabotype 1, 2, and 3) ^17^. Metabotype-specific differences in serum protein concentrations were found particularly for metabolic hormones, such as the increased FABP4 seen in both metabotype 1 and 2, whereas increased insulin and leptin were primarily observed in metabotype 2. The effects on immune system-related proteins generally remained relatively similar between the metabotype subgroups, such as reduced MPO and TGFα. To this end, two exceptions were CCL24, which expressed a specific increase in the metabotype 1 subgroup, and IL-8, which showed a specific decrease in the metabotype 3 subgroup.

**Figure 6.**
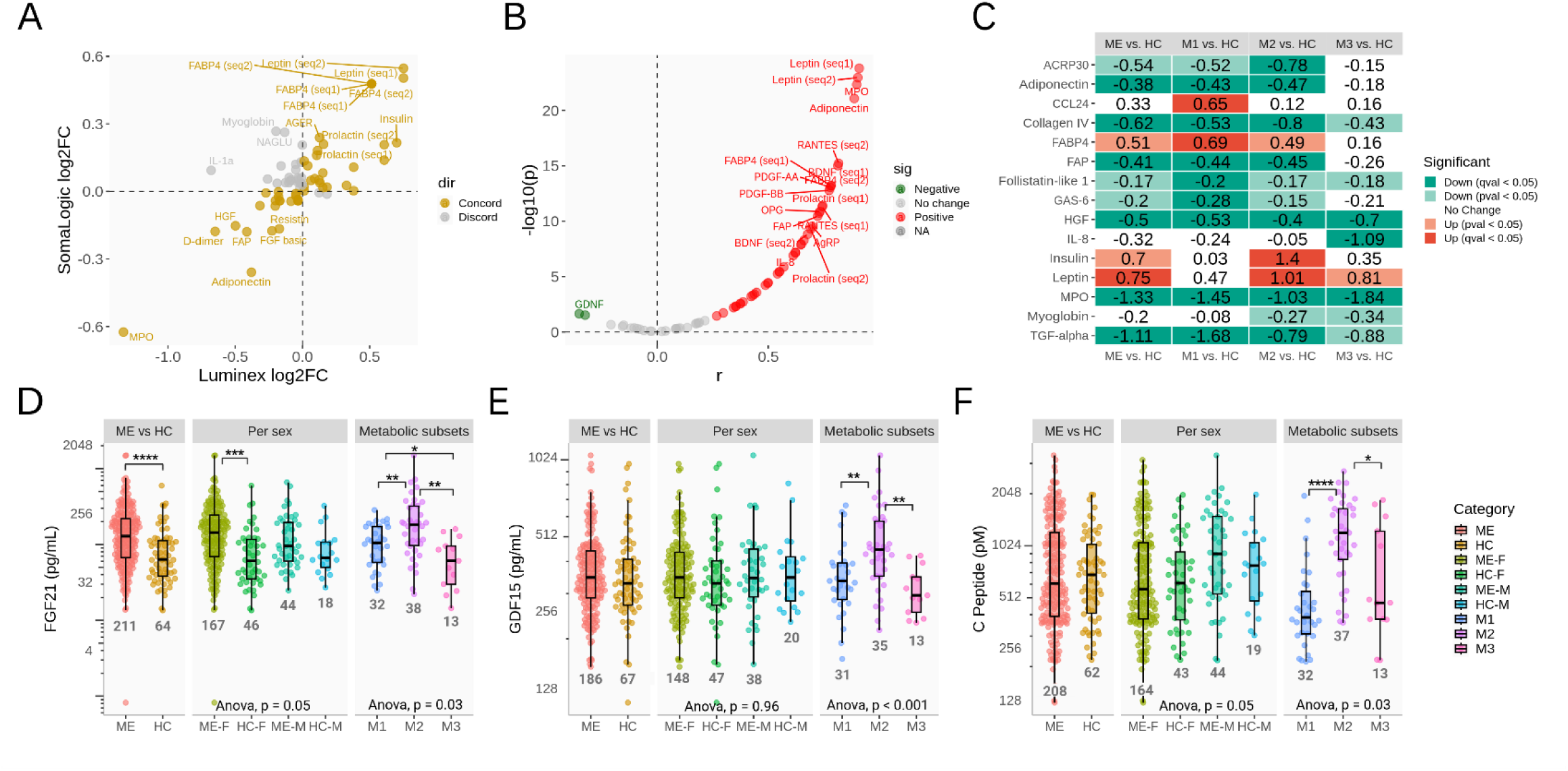
Targeted validation and expansion of key findings. Antibody-based measurements were used to validate and expand on key outcomes from aptamer-based serum proteomics. Antibody-based measurements (Luminex) were performed of a panel of 49 serum proteins related to immunity and metabolism, in 83 ME/CFS patient and 48 HC subjects. A) and B) show the correlation analysis between antibody-based and the aptamer-based analysis per protein. F) The heatmap display significantly affected proteins (antibody-detected), and comparison between the three ME/CFS metabotypes. C) Additional ELISA analysis of FGF-21, GDF15 and C-peptide was performed in larger cohorts (n=212 ME/CFS, n=66 HC).

We performed additional conventional ELISA measurements of two metabolic stress hormones, FGF-21 and GDF15, in addition to C-peptide, which indicate insulin production, using a larger cohort of subjects (n=212 ME/CFS, n=66 HC) (Figures 6D-F). FGF21 and C-peptide were not included in the previous aptamer-based analysis. For GDF15, the ELISA analysis showed no difference between the ME/CFS and HC groups, which supported the finding of the aptamer-based analysis (Figure 6E). For FGF-21, the serum concentration was higher in the ME/CFS group compared to the HC group (Figure 6D). When comparing by sex, the increase of FGF-21 was statistically significant in women, but not in men. The patients in the metabotype 2 subgroup had significantly higher serum concentrations of FGF-21 compared to the two other subgroups. While the serum concentrations of GDF15 and C-peptide were not significantly different between the ME/CFS and HC groups, irrespective of sex, also these showed higher levels in the metabotype 2 subgroup compared to metabotypes 1 and 3.

## Discussion

This serum proteomics study identified comprehensive differences in circulatory protein concentrations between ME/CFS patients and healthy individuals. Through the investigations of the data, we aimed to position the observed patterns of changes within the mechanistic landscape of tissue involvement and functional impact, as summarized in Figure 7.

**Figure 7.**
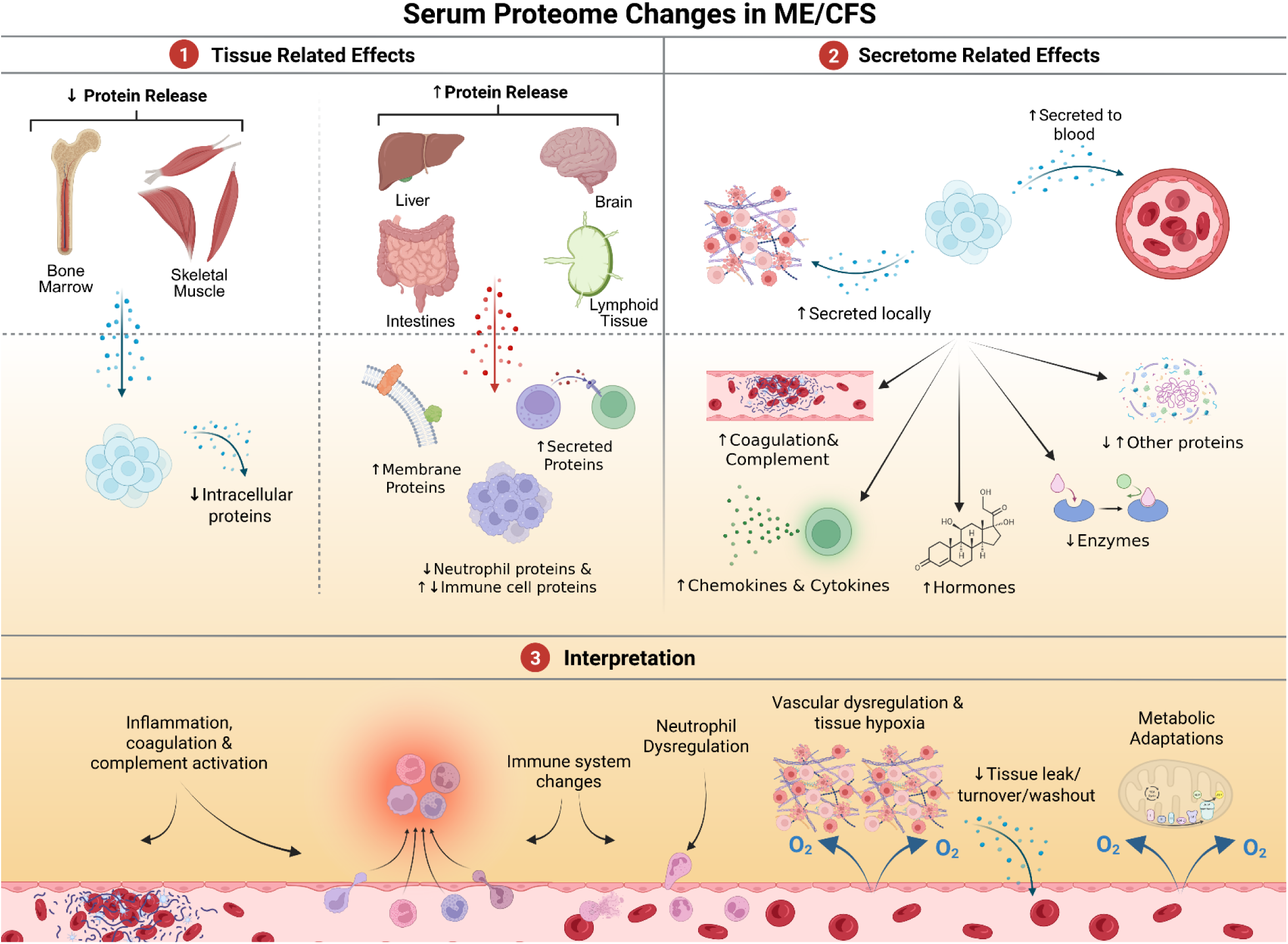
Impacts of serum proteome changes in ME/CFS. This illustration provides an overview of the key findings from our serum proteome study in ME/CFS, highlighting tissue-specific impacts, secretome alterations, and our interpretations: 1) Tissue-related effects: The analysis revealed distinct patterns for tissue-specific proteins from the brain, intestine, liver, skeletal muscle, and lymphoid tissue. ME/CFS patients showed increased levels of secreted proteins from these tissues, while intracellular proteins, particularly from skeletal muscle and bone marrow, were reduced. There was a specific reduction in the serum levels of proteins released from neutrophils, suggesting altered function of this cell type. 2) Secretome effects: The secretome analysis indicated an overall elevated state of secretory activity in ME/CFS patients. A pattern of increase was seen for proteins involved in coagulation and complement activation, chemokines, cytokines, and hormones, whereas enzymes showed a reduction. 3) Interpretation: The altered protein patterns support the notion of a root mechanism involving autoimmunity and/or immune dysregulation, and consequent roles of low-grade inflammation, tissue hypoperfusion, and metabolic alterations in ME/CFS.

We used two main approaches to uncover multiscale insights about the changes occurring in the body due to ME/CFS pathology. First, we analyzed the entire serum proteome dataset to assess systemic, tissue-specific, and immune cell-related impacts. Second, we focused on secreted proteins to examine changes in functional aspects such as signaling, transport, immune defense, and metabolism. We did thorough analyses to characterize the influence of sex, age, BMI, and fasting state, and it was confirmed that these covariates could not explain the changes observed in the ME/CFS group compared to the HC group. Furthermore, the comprehensive changes found to be unrelated to physical function argue against deconditioning as a driving factor for the findings in the ME/CFS group. However, this does not exclude the possibility that some of the covariates may influence biological mechanisms involved in symptom generation and maintenance, such as metabolic dysregulation. This is possibly exemplified by the worse physical function of the metabotype 2 subgroup ^17^ and its association with relatively high levels of insulin and leptin, combined with specific increases in metabolic stress hormones such as FGF21 and GDF15 (Figure 6).

### Reduced protein release from muscle

The widespread decrease of intracellular proteins released from skeletal muscle to blood may have different explanations. Possible systematic variation, such as dilution effects due to possible blood volume changes in ME/CFS ^47^, seems unlikely based on the specificity of the findings related to different tissues and processes. It could be speculated that reduced leakage of muscle proteins into the bloodstream could be associated with low physical activity and less muscle mass, but this is not convincingly supported since the data were adjusted for age, BMI, and sex, which indirectly integrates variation in muscle mass and activity. Yet, we did observe a reduction in COMP, a component of cartilage matrix that temporarily increases in serum upon physical exercise ^48^. Alternative explanations may involve aspects of skeletal muscle tissue maintenance and protein washout, and potentially, impaired tissue perfusion causing reduced protein flux from the interstitial fluid to blood. Given that different muscle proteins have different release pathways and clearance dynamics, it seems plausible that a pathomechanism involving vascular dysfunction and microvascular shunting could lead to reduced drainage of cellular protein remains from the tissue in ME/CFS patients. If this is true, the widespread reduction in intracellular muscle proteins in blood may be linked to muscle abnormality and key symptoms in ME/CFS, such as pain, fatigue and PEM, through circulatory and vascular dysfunction ^18,49–51^.

### Secreted proteins and their functional impact

The secretome provides an opportunity to interpret regulatory and functional impacts, as many of these proteins have established extracellular roles as signaling molecules, enzymes, structural proteins, transport proteins, and defense proteins. Our data indicated an elevated overall state of secretory activity in ME/CFS patients, with 62% of the affected secreted proteins showing increased concentrations. Many of these proteins were functionally annotated to coagulation, complement activation, chemokines, interleukins, hormones, enzyme inhibitors, and receptors. Increased blood levels of hormones and cytokines related to inflammation have been observed previously in individuals with ME/CFS, including IL-1, IL-6, IL-7, IL-17, and TNFalpha, although there is some variation between the available findings ^22,52,53^. Additionally, proteins not classified as secreted may act as circulatory messengers when present in blood, such as the tissue-specific FABP types, which function as fatty acid transporters inside cells and as circulating stress messengers when released from cells ^54^. In summary, the pattern of elevated secretory activity in the ME/CFS group is mechanistically consistent with immune dysregulation, inflammatory responses, and metabolic stress.

### Immune cell changes and attenuated neutrophil function

Comparison of immune cell-specific protein panels between ME/CFS patients and healthy controls suggested that multiple proteins involved in the functions and interactions of B-cells, dendritic cells, monocytes, NK-cells, and T-cells are affected by the disease. The most increased protein was malignant T cell-amplified sequence 1 (MCTS1), an oncogene originally found in human T-cell lymphoma that recently was identified as a potential biomarker for COVID-19-related thrombosis ^39^. Immune dysregulation, as reported in multiple studies ^15,16,55^, may be mechanistically associated with an underlying autoimmune pathomechanism, the overall state of elevated secretory activity, and increased levels of factors involved in inflammation, coagulation, and complement activity. Similar observations have previously been highlighted in relation to ME/CFS and long COVID ^15,56^. To this end, changes in coagulability, platelet hyperactivation, and fibrinaloid microclot formation has been found in ME/CFS, although not to the same extent as in Long COVID ^57^. A recent single-cell transcriptomics study of peripheral blood mononuclear cells (PBMCs, which does not include neutrophils) found both monocyte dysregulation and platelet hyperactivation in ME/CFS patients, although the abnormal platelet state was less pronounced 24 hours post-exercise ^58^.

Regarding the reduction of bone marrow proteins, decreased protein release from neutrophils appears to be a plausible explanation. Many of the granulocyte-related proteins found to be decreased in the patients are associated with neutrophil granules, stimulated secretion, and NETs, suggesting that changes in neutrophil maturation and/or activation states occur in ME/CFS ^59^. These cells release DNA and proteins such as MPO, BPI, histones, and granular proteins when activated upon infection. The release of these proteins is not restricted to the NET process, and other aspects of neutrophil activity and phenotypes may be involved ^60^. Since standard clinical blood cell counts did not indicate detectable abnormalities in the circulating numbers of the main leukocyte classes in the ME/CFS patients, the proteomics data may suggest that functional changes are present, which may include exhaustion or suppression of neutrophil activity. This represents a mechanistic element that also is likely to involve other immune cell types, as well as cells in the blood vessel walls ^61^.

### Selective increase in myoglobin

Based on the overall pattern of lower serum concentrations of many muscle-related proteins, the selective increase in myoglobin is of a certain interest, as it suggests muscle tissue oxygenation as a potential contributing factor. Nevertheless, as the increase in myoglobin was not reproduced by the antibody-based method, concern needs to be taken for the interpretation, and further investigations are required. Aptamer-based detection is considered a stable proteomics platform that is less susceptible to protein degradation^62^; however, it remains a semi-quantitative method with some limitations. Myoglobin in blood is a sensitive marker for muscle injury, but since the increase in ME/CFS patients was not accompanied by increases of other markers such as CKM or LDH, it does not seem to reflect tissue damage or differences in muscle mass in this case. In support, others have found reduced blood levels of creatine kinase in ME/CFS patients^25^. Instead, it may be speculated that the possible specific increase in myoglobin could be mechanistically associated with tissue hypoxia adaptation, serving as a compensation that contributes to improved muscle oxygenation. The primary function of myoglobin is to store and facilitate the transfer of oxygen from the cell membrane to the mitochondria inside muscle cells. This may align with our finding that serum from ME/CFS patients contain factors that seem to induce compensatory increases in mitochondrial respiration in muscle cells ^63^. Furthermore, this may involve elements of physiological mitigation that could be beneficial in resting ME/CFS patients, when blood lactate levels usually are within the normal range. However, upon exertion, the limitations become apparent, as indicated by the abnormal rise in lactate concentration at low workloads ^64^.

### Vascular dysregulation and hypoxia response

Several proteins associated with microcirculation and hypoxia response showed significant effects in ME/CFS patients. There was a reduction in VIM, a structural intracellular protein expressed by various cell types, which interacts with proteins such as CD44 on the endothelial surface ^34^. Reduced levels of ANTXR2, which mediates endothelial regulation ^35^, and CXADR, which mediates endothelial cell responses to shear stress ^36^, were also observed. Additionally, the reduced level of ATP5IF1 may indicate impaired protection against mitochondrial stress during oxygen deprivation ^37^. Regarding proteins that showed an increase in the patients, LIPG is an endothelial protein involved in lipoprotein metabolism and vascular function ^42^, which is released during immune activation^43^, and FAM20A, which has a possible function in hematopoiesis as secreted signaling protein ^65^. Several studies have documented endothelial dysfunction in ME/CFS patients ^66–68^. The endothelium forms the inner lining of blood vessels and plays important roles in the control of blood flow, immune reactions, and metabolism. Endothelial cell functions are regulated by circulating substances and interactions with other cells in the microenvironment, such as blood cells and smooth muscle cells ^69^. The contractile activity of the vessel wall (vascular tone) is a major determinant of blood flow through tissues, but the endothelial surface is also an important site of platelet and immune cell adhesion, aggregation, and activation. Oxygen-dependent mitochondrial ATP production is also a determinant of endothelial cell control of vascular tone ^70^, and red blood cell function and deformability ^71^. Capillary alterations in skeletal muscle and changes in hypoxia-related blood factors were recently reported in long COVID patients ^72,73^, and roles of immune dysfunction, inefficient oxygen transport and inflammatory disequilibrium have been proposed as drivers of persisting symptoms ^74^, which may be relevant for ME/CFS. This notion is further supported by the activation of the inherent hypoxia response in muscle tissue, as the expression of WASF3, a target gene for the HIF1A transcription factor, was increased in ME/CFS patients ^75^. Thus, impaired vascular flow and tissue hypoxia, with a role in endothelial dysfunction, may explain the interrelated patterns of metabolites and proteins that we observed in ME/CFS patients.

### Metabolic stress and energy metabolism

Our findings complement previous reports suggesting the involvement of serum proteins that signal metabolic stress in the context of ME/CFS, such as GDF-15 ^76^, FGF-21, resistin, leptin, and HGF ^22,52^. FGF-21 and GDF-15 are metabolic stress hormones, and increased levels have been found in conditions such as mitochondrial myopathies, type 2 diabetes, and hypoxia. Some hormones involved in metabolic adaptations showed associations with the previously described metabolic phenotypes in ME/CFS patients ^17^. For instance, based on the expanded targeted validation measurements, c-peptide (an insulin marker), leptin, and FGF21 were particularly increased in the metabotype 2 subset, whereas FABP4 only showed a significant increase in the metabotype 1 subset. In the normal healthy context, increased blood levels of so-called “exerkines,” including proteins such as FGF21 and GDF15, and other molecules such as lactate, free fatty acids (FFAs), and catecholamines, are associated with the physiological effects of exercise ^77^. These messengers act across multiple organ systems. Our findings in ME/CFS patients may align with a persistent state of elevated energy strain, which may reduce physical performance and prevent adequate restitution after exertion. Mechanisms causing tissue hypoperfusion could trigger of such metabolic stress ^78^, as supported by the link between reduced oxygen extraction and persistent exertional intolerance in metabolic myopathies ^79^, in ME/CFS ^80^, and after COVID-19 ^81^.

### Limitations of the study

There are some limitations of the study that should be considered when evaluating the findings. The aptamer technology provides measurement of relative protein levels, and findings of high interest should be verified using additional high-confidence methods. We found a reasonably good correlation between the aptamer-based proteomics analysis and the expanded analysis using antibody-based methods in our study. However, the study sample size is relatively small, although it represent one of the most comprehensive proteomics studies in this patient group. When using protein annotations to track functions and tissue locations, it is important to be aware that few proteins are exclusively specific to a given process or organ and may play different context-dependent roles. Therefore, alternative interpretations of the findings may be relevant. We have mitigated some of these limitations by primarily focusing our interpretations on patterns of changes related to different tissues and processes, rather than single protein effects. For some proteins, different aptamers detected different versions or domains of the same polypeptide. These may theoretically be utilized for purposes of internal validation or investigations of structural changes or peptide cleavage.

### Perspectives

There are several ways in which we believe this work could support new research directions. First, it identifies specific cells, organs, and processes affected in ME/CFS, which may guide the development of diagnostic markers and therapeutic strategies. These insights may also be relevant to related syndromes, such as long COVID and fibromyalgia. Second, it provides a mechanistic landscape that can be used to contextualize previous and future findings within a broader (patho)physiological framework - for example, in relation to potential risk factors identified through multi-omics analyses ^82,83^. Third, it may inform the development of relevant laboratory models for ME/CFS, including through systemic comparisons ^84^, the selection of appropriate cell types, and *in vitro* recapitulation of key mechanistic features ^63^.

### Conclusion

Our findings characterize a pathophysiology in ME/CFS composed of interrelated elements, including immune dysregulation, vascular dysfunction, metabolic stress, and chronic inflammation. The observed multiscale patterns of change converge on a mechanism involving coordinated immune, vascular, and metabolic disturbances. We hope these insights will support further research into disease mechanisms, therapeutic targets, and the development of predictive and diagnostic biomarkers, ultimately benefiting individuals affected by ME/CFS.

## Materials and methods

### Study cohort and samples

To investigate serum proteome changes in ME/CFS, we utilized 54 baseline ME/CFS patient samples from the two clinical intervention studies RituxME ^31^ and CycloME ^32^, and 27 sex- and age-matched healthy controls, randomly selected from the ME/CFS biobank at Haukeland University Hospital. Characteristics of the study cohort are provided in Table 1. The included healthy controls and 40 of the ME/CFS patients were non-fasting at the time of sample collection, and these patient had previously been assigned to three subgroups based on their metabolic phenotypes (metabotype 1, 2, and 3) ^17^. To facilitate evaluation of the impact of fasting, we included 10 additional patients who had performed overnight fasting before the sampling. In the validation measurements towards the end, samples were randomly picked from the ME/CFS biobank, including up to 212 ME/CFS patients and 66 healthy individuals. Clinical blood and serum analyses were performed according to standard laboratory routines at the hospital.

### Data processing and analysis

Serum samples were analyzed using the SomaScan platform (SomaLogic Operating Co., Inc.), yielding a dataset for 7326 specific aptamer-detected protein targets. Outlier detection removed four samples based on Mahalanobis distances of log10-transformed values, followed by PCA and a chi-square test (p < 0.1). High-leverage aptamers were identified using Z-scores (±1.8, corresponding to the 2.5th and 97.5th percentiles) and assigned NA, after which missing values were imputed using the variable’s minimum and maximum post-outlier removal. Given these adjustments, log10-transformed data were back-transformed using base-10 antilogarithms, followed by a final log2 transformation. Multivariate analysis involved PCA, with the first two principal components used for sample projections, and clinical variables correlated with sample coordinates via the envfit function (Vegan v2.4.0). Univariate analysis was performed using linear regression (adjusting for age, sex, BMI, and fasting baseline) with limma ^85^. For the purpose of hypothesis generation, we primarily referred to p-statistics to cover a wider spectrum of biological aspects potentially playing a role in ME/CFS. Explained variance was further assessed on variables with statistical significance (p < 0.05) using the function fitExtractVarPartModel in the VariancePartition library (GitHub - DiseaseNeuroGenomics/variancePartition). Semi-partial Pearson correlation coefficients were calculated for relevant subsets, adjusted for the same covariates, were calculated using ppcor ^86^, filtered by a standard cutoff (p < 0.05, absolute r > 0.3) visualized in a clinical-protein network, where protein communities were defined by shared clinical interactions. Functional enrichment was conducted using Gene Ontology ^87^ with ClusterProfiler (GitHub - YuLab-SMU/clusterProfiler), while ligand-receptor interactions were annotated via CellLinker ^88^, and functional/spatial annotations were retrieved from the Human Protein Atlas ^46,89^. All visualizations were generated using ggplot2, ComplexHeatmap, tidygraph, igraph, ggnet, and ggpubr. Data analysis was conducted in R ^90^ within the RStudio environment ^91^.

### Targeted immunodetection (ELISA and Luminex)

ELISA was performed using Quantikine Ready-to-Use ELISA kits (R&D Systems) for FGF-21 (Cat#: DF2100), C-peptide (Cat#: DICP00), and GDF-15 (Cat#: DG0150). The serum samples were diluted and measured in duplicates according to the manufacturer’s recommendations. Measurements were performed using the Spark microplate reader (Tecan Trading AG, Switzerland). Standard curve generation was conducted in Excel or GraphPad Prism, with further data analysis done in GraphPad Prism. Custom multiplex assays covering a total of 77 targets (SupplData9) were designed using the Luminex Assay Customization tool on the supplier’s website (Cat# LXSAMH, R&D Systems) and performed according to the manufacturer’s recommendations. Five custom assays were designed to cover the analytical targets, and two 96-well plates for each assay were used. The majority of the samples were analyzed in single reaction wells; however, some samples were analyzed as duplicates or included on multiple plates for quality control. Each plate included fresh target standard dilutions in duplicates. The serum samples were diluted according to the protocol provided by the manufacturer. Plates with identical assays were run on the same day. Measurements were performed using the Luminex 200 instrument (Luminex Corp.) and the accompanying analysis software to generate the standard curves and sample concentrations. Measurements that were beyond the range of the standard curve were excluded, and analytes that displayed an overweight of such measurements were also excluded from further analysis.

## Data Availability

All data produced in the present work are contained as supplementary files to the manuscript

## Resource availability

### Lead contact

Further information and requests for resources and reagents should be directed to and will be fulfilled by the lead contact, Karl Johan Tronstad

### Materials availability

This study did not generate new unique reagents.

### Data and code availability

- Analyzed proteomics data provided by the Somalogic is included as supplemental data.
- Any additional information required to reanalyze the data reported in this paper is available from the lead contact upon request.
- This paper does not generate original code.

## Acknowledgement

The authors thank the study personnel at each center of the “RituxME” and “CycloME” clinical trials for their efforts in patient follow-up and biobank sampling. This work received financial support from The Kavli Trust, the Research Council of Norway (KJT, projects 272680 and 343168), the Western Norway Regional Health Authority (Helse Vest) and the University of Bergen (PhD grants to FH and AH). The clinical trials that recruited patients for the biobank and the laboratory analyses in this study were financially supported by the Research Council of Norway, the Norwegian Regional Health Trusts, the MEandYou Foundation, the Norwegian ME Association, and the legacy of Torstein Hereid. Additionally, we are grateful for the contributions from private donors and the support from the Norwegian ME Association (Norges ME-forening).

## Author contributions

AH, FH, HC, ØF, OM, and KJT designed the analytical approach. ØF, OM, IGR, and KS included patients in clinical studies and provided biobank samples. KR, KA, FH, SD, and AH performed laboratory measurements. KAB and HPM provided scientific and technical advice. AH, FH, HC, KJT, and ØF conducted data analyses. AH, FH, HC, and KJT wrote the first version of manuscript. All authors approved the final version of the manuscript.

## Declaration of interests

The authors have declared that no conflict of interest exists.

## Declaration of generative AI and AI-assisted technologies

During the preparation of this manuscript, the author(s) used Microsoft Copilot (UiB institutional license) to assist with translation and ensure readability in certain sections.

## Supplemental information

- *DocumentS1:* Table S1 and Figure S1
- *SupplData1:* Excel file containing log2-transformed aptamer data, and subject metadata.
- *SupplData2:* Excel file containing univariate statistics, and cellular compartment metadata, comparing the ME/CFS group and the three metabotype subgroups, with the HC-group.
- *SupplData3:* Excel file containing PCA protein loading values, showing influence on PC1 and PC2.
- *SupplData4:* Excel file containing the percentage of variance explained by covariates.
- *SupplData5:* Excel file containing semi-partial Pearson correlation analysis between proteins and ME/CFS diagnosis, metabotype and physical function level (SF-36PF)
- *SupplData6:* Excel file containing univariate statistics for proteins in the MitoCarta 3.0 library.
- *SupplData7:* Excel file containing univariate statistics for tissue- and immune cell-specific proteins, including respective protein metadata.
- *SupplData8:* Excel file containing univariate statistics for the secretome, including protein function metadata; comparing the ME/CFS group and the three metabotype subgroups, with the HC-group.
- *SupplData9:* Excel file containing univariate statistics for the Luminex analysis; comparing the ME/CFS group and the three metabotype subgroups, with the HC-group.

## Notes

### Competing Interest Statement

The authors have declared no competing interest.

### Clinical Trial

NCT02229942, NCT02444091

### Author Declarations

The Regional Committees for Medical and Health Research Ethics in Norway gave ethical approval for the work.

