## Supplementary material for "Charting the Circulating Proteome in ME/CFS: Cross System Profiling and Mechanistic insights": DocumentS1.docx

**Supplementary information**

**Table S1 – Blood cell data for the ME/CFS group**

| Measurement | Mean | SD | n | Normal | Unit |
| --- | --- | --- | --- | --- | --- |
| Hemoglobin | 14.26 | 1.16 | 41 | *11.7 - 15.3* | g/dL |
| MCV | 88.54 | 2.78 | 41 | *82 - 98* | fL |
| Leucocytes | 6.23 | 1.35 | 41 | *4 - 10* | 10^9^/L |
| Neutrophile granulocytes | 3.38 | 1.03 | 41 | *1.5 - 7.3* | 10^9^/L |
| Lymphocytes | 2.20 | 0.80 | 41 | *1.1 - 3.3* | 10^9^/L |
| Monocytes | 0.45 | 0.12 | 41 | *0.2 - 0.8* | 10^9^/L |
| Eosinophiles | 0.16 | 0.11 | 41 | *< 0.4* | 10^9^/L |
| Basofiles | 0.05 | 0.05 | 41 | *< 0.2* | 10^9^/L |
| Thrombocytes | 273.49 | 64.39 | 41 | *145 - 350* | 10^9^/L |

Summary of the data available from the ME/CFS patients. The ME/CFS samples were obtained from participants in the RituxME and CycloME trials. Measurements were performed in the clinic.

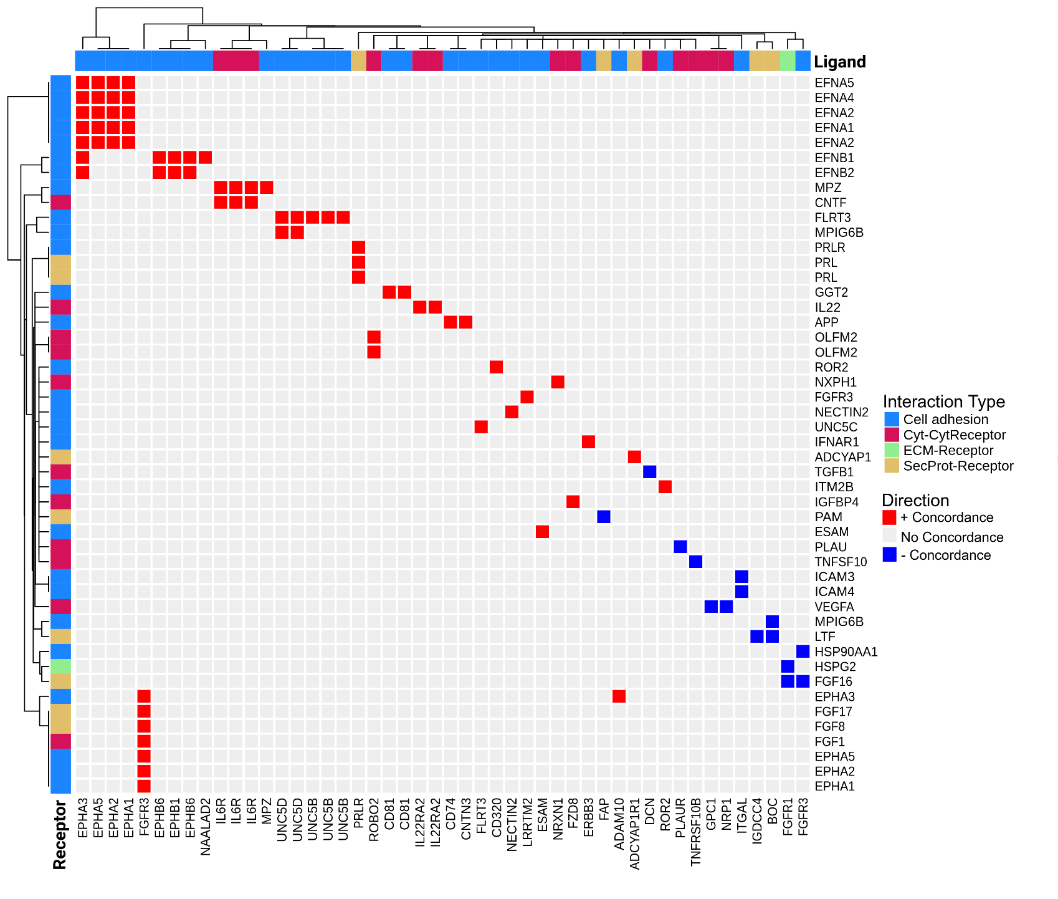

**Figure S1: Ligand – receptor interactions**

We performed a ligand - receptor interaction analysis to attempt elucidating possible patterns of regulation that could be related to the pathomechanisms of ME/CFS
